# Predicting Atrial Fibrillation Recurrence After Electrical Cardioversion

**DOI:** 10.1101/2025.09.29.25336937

**Authors:** Sittinun Thangjui, Emmett Helsel, Efeturi Okorigba, Anh Q Le, Saim Rana, Fatima Asad, Emily Hendricks, Sandeep Arora, David Schwartzman, Sudarshan Balla

**Affiliations:** Department of Cardiology, Heart and Vascular Institute, West Virginia University, Morgantown, WV; Department of Internal Medicine, West Virginia University, Morgantown, WV; Section of Electrophysiology, Department of Cardiology, Heart and Vascular Institute, West Virginia University, Morgantown, WV

**Keywords:** Atrial fibrillation, left atrial volume index, cardioversion, antiarrhythmic drugs

## Abstract

**Background:** Predicting atrial fibrillation (AF) recurrence after external cardioversion (ECV) is challenging. The SLAC score predicts 6-month recurrence but lacked external validation. We aimed to externally validate the SLAC score and develop an improved predictive model.

**Methods:** We conducted a single-center retrospective study of patients who underwent successful ECV for AF between 2015 and 2020. Patients with atrial flutter, complex congenital heart disease or early AF ablation were excluded. The primary outcome was AF recurrence at 6 months. SLAC score performance was tested, and multivariable logistic regression was used to develop the SLASH score. Discrimination was assessed with the area under the curve (AUC), calibration with Hosmer–Lemeshow testing, and internal validation with bootstrapping.

**Results:** Of 361 patients (mean age 66 ± 12 years, 61% male), 53.7% experienced AF recurrence in 6 months. Median SLAC scores were higher in recurrence patients (7 vs 2). The SLAC score demonstrated moderate discrimination (AUC 0.701), improved with a cutoff ≥6 (AUC 0.720). Independent predictors of recurrence included left atrial volume index ≥40 ml/m², prior stroke/TIA, heart rate > 70 bpm, smoking history, and absence of post-ECV antiarrhythmic therapy. These were incorporated into the SLASH score (0–8 points), which demonstrated improved discrimination (AUC 0.781). Recurrence rates were 16.1%, 65.4%, and 78.5% in low-, moderate-, and high-risk groups, respectively, with acceptable calibration (Hosmer–Lemeshow p=0.18).

**Conclusion:** The SLAC score provides moderate prediction of 6-month AF recurrence after ECV. The SLASH score showed superior performance and may improve risk stratification and rhythm management. Prospective validation is warranted.

## Introduction

The management of atrial fibrillation (AF) employs a multifaceted approach, with an equipoise between rhythm and rate control, which has endured for nearly two decades. However, the results of the Early Treatment of Atrial Fibrillation for Stroke Prevention Trial (EAST-AFNET4) trial demonstrated the superiority of early rhythm control compared to a rate control strategy in reducing adverse cardiovascular outcomes over 5 years in patients with early-stage AF (1). The most effective intervention to restore sinus rhythm is direct current external cardioversion (ECV), which immediately restores sinus rhythm and has been shown to improve quality of life within the first 3 months (2). However, cardioversion alone is often insufficient as up to 70% of patients experience AF recurrence after ECV (3). Additional strategies, such as antiarrhythmic therapy, catheter ablation, or surgical ablation, are often necessary to maintain a sinus rhythm, although none of these approaches is curative (1). While these treatments are generally safe, they carry risks of medication-related adverse effects, procedure-specific complications, and limited access, particularly in non-urban settings (1, 4). Approximately 40% of patients had a history of ECV in the EAST AFNET4 trial. Sinus rhythm was found more often in patients who had been randomly assigned to receive early rhythm control (82.1% at 2 years) than in patients assigned to receive usual care (60.5% at 2 years), suggesting the importance of therapies aimed at preventing recurrence and maintaining sinus rhythm. Consequently, a key clinical challenge is predicting AF recurrence in those who undergo ECV to better triage patients for additional approaches to maintain sinus rhythm.

Several risk factors are associated with an increased AF recurrence after cardioversion, including older age, female sex, previous cardioversion, chronic obstructive pulmonary disease (COPD), renal impairment, structural heart disease, and left atrial enlargement, especially measured by left atrial volume index (LAVI) (5–7). The previously published SLAC score predicts AF recurrence at 6 months following ECV (6). The components of the SLAC score included: history of stroke (3 points), LAVI ≥ 40 mL/m² (6 points), type of AF (persistent: 1 point; longstanding persistent: 4 points), and prior cardioversion (1 point). In the derivation cohort, the SLAC score demonstrated good discrimination, with an area under the ROC (AUC) of 0.739 (6).

One external validation study reported only fair discrimination, with an AUC of 0.602 (7). Notably, that study applied a simplified cut-off of 2 rather than the original three-level risk stratification, which may have limited the performance of the score. In this study, we aim to externally validate the SLAC score and investigate a prediction model with improved performance. Our goal is to provide a practical tool for clinicians to risk-stratify AF patients undergoing ECV and optimize individualized rhythm management strategies post-ECV.

## Methods

We performed a single center retrospective cohort study to validate the previously published SLAC score for predicting AF recurrence after ECV. Data were obtained from a review of electronic medical records, and the study protocol was approved by the West Virginia University (WVU) Institutional Review Board (protocol #2309847870). The study was reported in accordance with the STROBE (Strengthening the Reporting of Observational Studies in Epidemiology) checklist for cohort studies (8).

Patients with a confirmed diagnosis of AF (paroxysmal, persistent, long-standing persistent) who underwent successful external electrical cardioversion between January 1, 2015, and December 31, 2020, in the WVU Health System were included. Inclusion criteria included a transthoracic echocardiogram performed 6 months before or 1 month after ECV, successful restoration of sinus rhythm, and reported heart rhythm at 6-month follow-up. Patients with atrial flutter, cardioversion performed during open-heart surgery, surgical or catheter AF ablation within 6 months after the cardioversion, or complex congenital heart disease were excluded. The primary outcome was recurrence of AF within 6 months post ECV.

Baseline clinical, laboratory, and procedural data were collected, including demographics, comorbidities, AF type, medications, clinical scores (CHA₂DS₂-VASc, HAS-BLED, SLAC), and echocardiographic parameters. Echocardiograms were performed using standard systems, Philips Epiq 7 (Philips Medical Systems, Andover, Massachusetts) or GE Vivid E95 (GE Healthcare, Chicago, Illinois) systems. Left atrial volume was derived from report if readily available or measured and interpreted according to the ASE guidelines (9). Electrical cardioversion was performed according to local institutional practice using biphasic energy with a Medtronic Lifepak 20 defibrillator (Medtronic, Minneapolis, MN); the delivered energy level and pad placement were at the discretion of the treating physician. Resting heart rate was obtained from a 12-lead electrocardiogram performed prior to ECV.

The derivation of the SLAC score has been reported previously (6). For the validation of SLAC score, we estimated that a total of 345 patients (assuming 50% recurrence rate) would be needed to provide a 95% CI ± 0.05 around the originally reported AUC of 0.739. Therefore, we aimed to include up to 400 patients to account for exclusions and missing data. Categorical variables were reported as frequencies (%) and compared with chi-square or Fisher’s exact test. Continuous variables with a normal distribution were expressed as mean ± SD, while non-normally distributed variables were expressed as median (IQR). Comparisons were made using the t-test and Wilcoxon rank-sum test, as appropriate. Logistic regression was used for univariable screening.

Development of an updated score was a secondary aim. Following TRIPOD guidance, we applied the 10-events-per-variable rule, with the number of predictors limited by total events (10). Variables statistically associated with AF recurrence in univariable analysis, along with the original SLAC score variables, were entered into a multivariable logistic regression model. Non-significant variables were sequentially removed, and regression coefficients from the final model were used to assign points for the score, with 1 point corresponding to the smallest coefficient and serving as the denominator for weighing the remaining variables. Backward elimination was applied, and model performance was assessed with AUC, diagnostic odds ratios, and Hosmer– Lemeshow testing. Internal validation was performed with 1,000 bootstrap samples. Discrimination was assessed using ROC curve analysis with AUC and 95% CI. Predictive accuracy was evaluated by calculating AF recurrence rates across low, moderate, and high-risk categories. The updated model was compared against the SLAC and ATLAS scores, and diagnostic performance, including sensitivity, specificity, and positive and negative likelihood ratios, was reported. We also constructed Kaplan–Meier survival curves to visualize AF recurrence at 6 months stratified by SLAC score risk groups (low, intermediate, and high). Differences between groups were assessed using the log-rank test. All analyses were conducted in STATA 19 (StataCorp, College Station, TX).

## Results

Of 1,045 patients undergoing electrical cardioversion for AF, 684 were excluded (AF ablation during follow-up n=53, atrial flutter n=55, duplicate records n=10, complex congenital heart disease n=8, perioperative cardioversion of cardiac surgery n=12, failed cardioversion n=20, incomplete/missing follow-up n=75, no cardioversion performed n=5, missing LAVI n=34, and unavailable/old echocardiogram n=412). The final cohort included 361 patients (mean age 65.9 ± 12.5 years, 39.0% female, 98.3% Caucasian), see Supplementary Figure 1 for study flow chart.

At 6 months, AF recurrence occurred in 53.7% (n=194) of patients. Compared with those maintaining sinus rhythm, patients with recurrence more often had a history of stroke (10.4% vs 4.8%, OR 2.30 [95%CI: 0.98–5.36], p=0.05), were smokers (51.8% vs 41.0%, OR 1.33 [95%CI: 1.06–1.66], p=0.01), and received lower rates of post-cardioversion antiarrhythmic therapy (43.5% vs 64.5%, OR 0.44 [95%CI: 0.29–0.68], p<0.01). Heart rate ≤70 beats per minute(bpm) before cardioversion was protective (13.4% vs 23.4%, OR 0.54 [95%CI: 0.31–0.94], p=0.03). Among echocardiographic parameters, only LAVI predicted recurrence (≥40 ml/m²: 75.8% vs 32.3%, OR 6.54 [95%CI: 4.13–10.38], p<0.01). Recurrence was also associated with higher mean biphasic shock energy (241.9 ± 81.7 vs 223.3 ± 74.9 J, p=0.03). Table 1 shows the baseline characteristics of the study population (Detailed baseline characteristics data are available in supplementary Table 1).

**Table 1.** Baseline characteristics of the study groups.

| Baseline characteristics | AF recurrence group (194) |  | Sinus rhythm group (167) |  | Odds ratio (95%CI) | p-value |
| --- | --- | --- | --- | --- | --- | --- |
|  | n | % | n | % |  |  |
| Demographic |  |  |  |  |  |  |
| Age, years (mean ± SD) | 66.79 | 11.43 | 65.04 | 13.29 | 1.01 (0.99-1.03) | 0.18 |
| Female sex | 89 | 45.88 | 52 | 31.14 | 1.87 (1.22-2.89) | 0.04 |
| Race |  |  |  |  |  |  |
| • Caucasian | 190 | 97.94 | 165 | 98.8 | 1.28 (0.39-4.18) | 0.69 |
| • Black | 3 | 1.55 | 1 | 0.60 | 2.61 (0.27-25.29) | 0.41 |
| Co-morbidities |  |  |  |  |  |  |
| Type of AF |  |  |  |  |  |  |
| • Paroxysmal AF | 83 | 42.78 | 68 | 40.72 | 1.09 (0.72-1.66) | 0.69 |
| • Persistent AF | 95 | 48.97 | 84 | 50.30 | 0.93 (0.60-1.43) | 0.73 |
| • Long-standing persistent AF | 16 | 8.25 | 15 | 3.98 | 0.87 (0.40-1.89) | 0.73 |
| Hypertension | 168 | 86.60 | 145 | 86.83 | 0.98 (0.53-1.80) | 0.95 |
| Previous cardioversion | 47 | 24.23 | 46 | 27.54 | 0.84 (0.52-1.35) | 0.47 |
| Coronary artery disease | 59 | 30.41 | 55 | 32.93 | 0.89 (0.57-1.39) | 0.61 |
| Diabetes mellitus | 57 | 29.38 | 54 | 32.34 | 0.87 (0.56-1.36) | 0.54 |
| Hyperlipidemia | 139 | 71.65 | 129 | 77.25 | 0.74 (0.46-1.20) | 0.23 |
| Stroke/TIA | 20 | 10.36 | 8 | 4.79 | 2.30 (0.98-5.36) | 0.05 |
| Peripheral artery disease | 15 | 7.77 | 6 | 3.61 | 2.25 (0.85-5.93) | 0.10 |
| COPD | 38 | 19.69 | 31 | 18.56 | 1.08 (0.63-1.82) | 0.79 |
| Obesity | 131 | 67.53 | 108 | 64.67 | 1.14 (0.73-1.76) | 0.57 |
| CKD stage 3 and above | 39 | 20.21 | 28 | 16.87 | 1.25 (0.73-2.14) | 0.42 |
| Acute decompensated HF | 20 | 10.31 | 16 | 9.58 | 1.08 (0.54-2.17) | 0.82 |
| Heart failure (HF) |  |  |  |  |  |  |
| • HF reduced ejection fraction | 46 | 23.71 | 43 | 25.75 | 0.93 (0.55-1.56) | 0.78 |
| • HF mildly reduced ejection fraction | 29 | 14.95 | 26 | 15.57 | 0.97 (0.52-1.79) | 0.92 |
| • HF preserved ejection fraction | 36 | 18.56 | 26 | 15.57 | 1.20 (0.66-2.18) | 0.55 |
| Valvular heart disease (≥ moderate severity) | 55 | 28.35 | 40 | 23.95 | 1.26 (0.78-2.01) | 0.34 |
| • Mitral regurgitation | 42 | 21.65 | 21 | 12.57 | 1.92 (1.09-3.40) | 0.03 |
| Valve replacement | 12 | 6.19 | 17 | 10.18 | 0.59 (0.27-1.26) | 0.17 |
| History of MAZE | 10 | 5.18 | 7 | 4.19 | 1.24 (0.46-3.36) | 0.66 |
| History of atrial fibrillation ablation | 14 | 7.25 | 14 | 8.38 | 0.85 (0.40-1.85) | 0.69 |
| History of left atrial appendage occlusion | 15 | 7.77 | 9 | 5.39 | 1.48 (0.63-3.45) | 0.37 |
| Previous cardioversion | 47 | 24.23 | 46 | 27.54 | 0.84 (0.52-1.35) | 0.47 |
| Obstructive sleep apnea | 46 | 25.14 | 53 | 33.97 | 0.77 (0.41-1.44) | 0.43 |
| History of smoking | 100 | 51.81 | 68 | 40.96 | 1.33 (1.06-1.66) | 0.01 |
| Alcohol use | 59 | 30.41 | 56 | 33.53 | 0.87 (0.56-1.35) | 0.53 |
| Risk score |  |  |  |  |  |  |
| CHA2DS2-VASc (median, IQR1-3) | 3 | 2-4 | 3 | 2-4 | 1.13 (0.99-1.29) | 0.07 |
| HAS-BLED, (median, IQR1-3) | 1 | 1-2 | 1 | 1-2 | 1.16 (0.93-1.44) | 0.19 |
| SLAC score (median, IQR1-3) | 7 | 6-7 | 2 | 1-6 | 1.28 (1.20-1.37) | < 0.01 |
| • Low risk (0-2 points) | 42 | 21.65 | 99 | 59.28 | 0.19 (0.12-0.30) | < 0.01 |
| • Moderate risk (3-7 points) | 104 | 53.61 | 52 | 31.14 | 2.56 (1.66-3.94) | < 0.01 |
| • High risk (8-14 points) | 48 | 24.74 | 16 | 9.58 | 3.10 (1.69-5.71) | < 0.01 |
| Physical exam |  |  |  |  |  |  |
| BMI, kg/m<sup>2</sup> (mean ± SD) | 33.70 | 7.78 | 33.89 | 8.29 | 0.99 (0.97-1.02) | 0.82 |
| Systolic BP, mmHg (mean ± SD) | 127.48 | 19.65 | 125.38 | 19.63 | 1.01 (0.99-1.02) | 0.31 |
| Diastolic BP, mmHg (mean ± SD) | 83.12 | 12.86 | 82.59 | 13.57 | 1.00 (0.99-1.02) | 0.70 |
| Heart rate, beat per minute (mean ± SD) | 94.17 | 25.59 | 90.32 | 23.96 | 1.01 (0.99-1.01) | 0.14 |
| • Heart rate > 70 beat per minute | 168 | 56.76 | 26 | 40.00 | 1.97 (1.14-3.40) | 0.02 |

| Medication |  |  |  |  |  |  |
| --- | --- | --- | --- | --- | --- | --- |
| Beta blocker | 151 | 77.84 | 134 | 80.24 | 0.86 (0.52-1.44) | 0.58 |
| Non-dihydropyridine CCB | 39 | 20.10 | 31 | 18.56 | 1.10 (0.65-1.87) | 0.71 |
| Antiarrhythmic medication |  |  |  |  |  |  |
| • None | 127 | 65.46 | 97 | 58.08 | 1.05 (0.58-1.88) | 0.88 |
| • Amiodarone | 48 | 24.74 | 55 | 32.93 | 0.67 (0.41-1.07) | 0.09 |
| • Sotalol/Dofetilide | 14 | 7.22 | 5 | 2.99 | 2.19 (0.74-6.14) | 0.16 |
| • Flecainide | 5 | 2.58 | 6 | 3.59 | 0.64 (0.19-2.15) | 0.47 |
| Digoxin | 22 | 11.34 | 22 | 13.17 | 0.84 (0.45-1.58) | 0.59 |
| Statin | 104 | 53.89 | 94 | 56.29 | 0.91 (0.60-1.38) | 0.65 |
| ACEI/ARB/ARNI | 103 | 53.09 | 90 | 53.89 | 0.97 (0.64-1.45) | 0.88 |
| Loop diuretics | 77 | 39.69 | 71 | 42.51 | 0.89 (0.58-1.36) | 0.59 |
| IV diuretics | 20 | 10.36 | 12 | 7.19 | 1.49 (0.71-3.15) | 0.29 |
| IV rate/rhythm control | 42 | 21.65 | 31 | 18.56 | 1.17 (0.91-1.51) | 0.21 |
| • Amiodarone | 6 | 3.09 | 9 | 5.39 | 0.60 (0.21-1.72) | 0.34 |
| • Calcium channel blocker | 26 | 13.40 | 18 | 10.78 | 1.29 (0.68-2.46) | 0.44 |
| • Beta blocker | 10 | 5.15 | 4 | 2.40 | 2.24 (0.69-7.30) | 0.18 |
| Anticoagulation prior to cardioversion | 160 | 82.47 | 139 | 83.23 | 0.95 (0.55-1.64) | 0.85 |
| Aspirin | 68 | 35.05 | 69 | 41.32 | 0.77 (0.50-1.17) | 0.22 |
| TTE parameters |  |  |  |  |  |  |
| LAVI group (mean ± SD) | 45.76 | 12.27 | 38.12 | 10.86 | 1.06 (1.04-1.09) | < 0.01 |
| • < 34 ml/m<sup>2</sup> | 31 | 15.98 | 64 | 38.55 | 0.30 (0.18-0.50) | < 0.01 |
| • 34-40 ml/m<sup>2</sup> | 16 | 8.25 | 48 | 28.92 | 0.22 (0.12-0.41) | < 0.01 |
| • 40-48 ml/m<sup>2</sup> | 73 | 37.63 | 44 | 37.6 | 3.11 (1.88-5.14) | < 0.01 |
| • ≥ 48 ml/m<sup>2</sup> | 74 | 38.14 | 27 | 16.27 | 3.17 (1.92-5.25) | < 0.01 |
| LAVI ≥ 40 ml/m<sup>2</sup> | 147 | 75.77 | 54 | 32.34 | 6.54 (4.13-10.38) | < 0.01 |
| LVEF | 50.08 | 15.32 | 49.39 | 15.46 | 1.00 (0.99-1.01) | 0.69 |
| • < 25 % | 18 | 9.28 | 15 | 8.98 | 1.04 (0.51-2.13) | 0.92 |
| • 25-40 % | 28 | 14.43 | 28 | 16.77 | 0.84 (0.47-1.48) | 0.54 |
| • 40-50 % | 29 | 14.95 | 26 | 15.57 | 0.95 (0.54-1.69) | 0.87 |
| • >50 % | 119 | 61.34 | 98 | 58.68 | 1.12 (0.73-1.70) | 0.61 |
| Right atrium area (cm<sup>2</sup>) (mean ± SD) | 22.28 | 5.63 | 21.15 | 5.71 | 1.04 (1.00-1.08) | 0.07 |
| Cardioversion procedure |  |  |  |  |  |  |
| Average biphasic energy, Joules (mean ± SD) | 241.93 | 81.72 | 223.32 | 74.91 | 1.00 (0.99-1.00) | 0.03 |
| Post-cardioversion bradycardia or hypotension | 9 | 4.81 | 11 | 6.75 | 0.70 (0.28-1.73) | 0.44 |
| Post cardioversion antiarrhythmics | 84 | 43.52 | 106 | 63.47 | 0.44 (0.29-0.68) | < 0.01 |
| • None | 109 | 64.12 | 61 | 36.53 | 2.25 (1.21-1.85) | < 0.01 |
| • Amiodarone | 61 | 31.61 | 79 | 47.31 | 0.43 (0.27-0.68) | < 0.01 |
| • Sotalol/Dofetilide | 15 | 7.77 | 12 | 7.19 | 0.70 (0.31-1.59) | 0.13 |
| • Flecainide or propafenone | 8 | 4.15 | 12 | 7.19 | 0.37 (0.14-0.96) | 0.04 |
| Follow up clinical outcomes at 6 months |  |  |  |  |  |  |
| Death | 1 | 1.35 | 0 | 0 | N/A | 0.31 |
| Pacemaker insertion | 4 | 2.07 | 4 | 2.52 | 0.82 (0.20-3.33) | 0.78 |
| Antiarrhythmic use | 91 | 46.91 | 82 | 49.40 | 0.96 (0.78-1.18) | 0.70 |

Abbreviation: ACEI = Angiotensin-converting-enzyme inhibitors; AF = atrial fibrillation; ARB = angiotensin II receptor blocker; ARNI = Angiotensin receptor neprilysin inhibitor; BMI = body mass index; BSA = body surface area; CCB = calcium channel blocker; CKD = chronic kidney disease; COPD = chronic obstructive pulmonary disease; g = gram; GLP1A = glucagon-like peptide-1 receptor agonists; HF = heart failure; kg = kilogram; LA = left atrium; LAVI = left atrial volume index; LVEF = left ventricular ejection fraction; ml = milliliter; m<sup>2</sup> = square meter; SD = standard deviation; SGLT2i = sodium-glucose cotransporter 2 inhibitors; TAPSE = tricuspid annular plane systolic excursion; TIA = transient ischemic attack; TTE = transthoracic echocardiography

The median SLAC score was higher in the AF recurrence group (7 [IQR 6–7]) than in the sinus rhythm group (2 [IQR 1–6]). The recurrence rates of 21.6%, 53.6%, and 75.8% were noted across low (SLAC score 0-2), moderate (SLAC score 3-7), and high-risk strata (SLAC score 8-14), respectively (p<0.01), see Figure 1A. The SLAC score showed fair discrimination (AUC 0.701), while a simplified cutoff ≥6 improved performances (sensitivity 72.1%, specificity 71.2%, AUC 0.720) as shown in Table 4.

**Figure 1.**
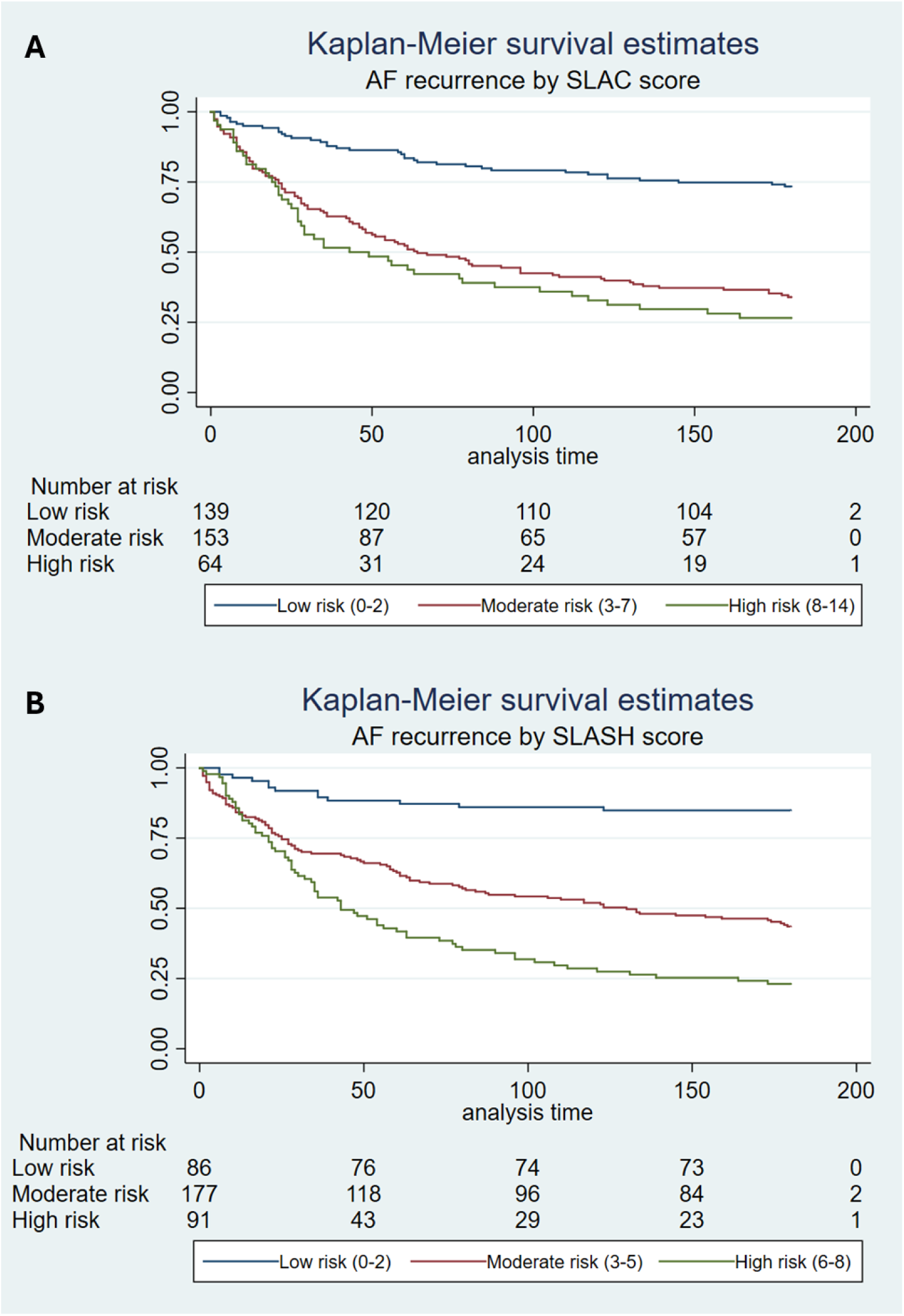
Kapla-Meier survival estimates stratified by SLAC and SLASH score Abbreviation: AF = atrial fibrillation

For new model development, variables with p < 0.1 in univariable analysis were considered. Backward multivariable logistic regression identified five independent predictors: LAVI ≥40 ml/m² (aOR 7.62 [95%CI: 4.63–12.53]), lack of antiarrhythmic drug therapy use post-cardioversion (aOR 2.53 [95%CI: 1.54–4.16]), history of smoking (aOR 1.58 [95%CI: 0.98–2.56]), heart rate >70 bpm (aOR 1.80 [95%CI: 0.95–3.43]), and history of stroke/TIA (aOR 2.17 [95%CI: 0.82–5.71]), see Table 2. We incorporated these variables into the new prediction model-**SLASH score** (**<u>S</u>**troke/TIA 1 point, **<u>L</u>**AVI ≥40 ml/m² 3 points, no **<u>A</u>**ntiarrhythmic therapy 2 points, **<u>S</u>**moking 1 point, **<u>H</u>**R >70 bpm 1 point; total 0–8). The history of stroke was retained in the model despite not reaching statistical significance because of its relatively high odds of recurrence and limited sample size; removing this variable negatively affected the overall predictive performance of the score, as shown in Table 3. In the internal cross-validation, the updated SLASH score demonstrated good discrimination (AUC 0.781 [95%CI: 0.73–0.83]) with simplified risk groups: low (0–2), moderate (3–5), and high (6–8). The simplified categorical version retained reasonable accuracy (AUC 0.734 [95%CI: 0.69–0.78]) with recurrence rates of 16.1%, 65.4%, and 78.5%, respectively (see Figure 1B). Calibration was acceptable (Hosmer– Lemeshow p=0.18), and internal bootstrapping confirmed discrimination (AUC 0.78 [95%CI: 0.73–0.82]).

**Table 2.** Multivariable logistic regression analysis showing the SLAC score model and additional independent variables in predicting atrial fibrillation recurrence at 6 months.

| Variables | Univariable OR | p-value | Multivariable OR | p-value | New reduced model multivariable OR | p-value |
| --- | --- | --- | --- | --- | --- | --- |
| Original SLAC score variables |  |  |  |  |  |  |
| Stroke/TIA | 2.30 (0.98-5.36) | 0.05 | 1.92 (0.70-5.29) | 0.21 | 2.17 (0.82-5.71) | 0.12 |
| LAVI cut point $\geq 40$ ml/m<sup>2</sup> | 6.54 (4.13-10.38) | <0.01 | 6.89 (4.10 – 11.59) | <0.01 | 7.62 (4.63-12.53) | < 0.01 |
| Type of AF |  |  |  |  |  |  |
| • Paroxysmal AF | 1.09 (0.72-1.66) | 0.69 | Reference | Reference |  |  |
| • Persistent AF | 0.93 (0.60-1.43) | 0.73 | 0.80 (0.49 – 1.35) | 0.41 |  |  |
| • Long-standing persistent AF | 0.87 (0.40-1.89) | 0.73 | 0.71 (0.29-1.74) | 0.45 |  |  |
| Previous cardioversion | 0.84 (0.52-1.35) | 0.47 | 1.11 (0.63-1.97) | 0.71 |  |  |
| Additional independent variable |  |  |  |  |  |  |
| History of smoking | 1.33 (1.06-1.66) | 0.01 | 1.46 (0.90-2.39) | 0.13 | 1.58 (0.98-2.56) | 0.06 |
| Heart rate > 70 beats per minute before cardioversion | 1.97 (1.14-3.40) | 0.02 | 1.76 (0.92-3.38) | 0.09 | 1.80 (0.95-3.43) | 0.07 |
| Peripheral artery disease | 2.25 (0.85-5.93) | 0.10 | 1.74 (0.58-5.22) | 0.33 |  |  |
| Lack of antiarrhythmic drug therapy after cardioversion | 2.25 (1.47-3.45) | <0.01 | 2.46 (1.46-4.15) | <0.01 | 2.53 (1.54 – 4.16) | <0.01 |
| Female | 1.87 (1.22-2.89) | 0.04 | 1.30 (0.76-2.23) | 0.34 |  |  |
| Right atrium area $\geq 24$ cm<sup>2</sup> | 1.44 (0.94-2.21) | 0.09 | 1.19 (0.71-2.01) | 0.50 | | |
| CHA2DS2-VASc $\geq 4$ | 1.54 (1.01-2.34) | 0.05 | 1.13 (0.65-1.98) | 0.66 | | |

Abbreviation: AF = atrial fibrillation; LAVI = left atrial volume index; ml/m<sup>2</sup> = milliliter/square meter; OR = odd ratio; TIA = transient ischemic attack

**Table 3.**
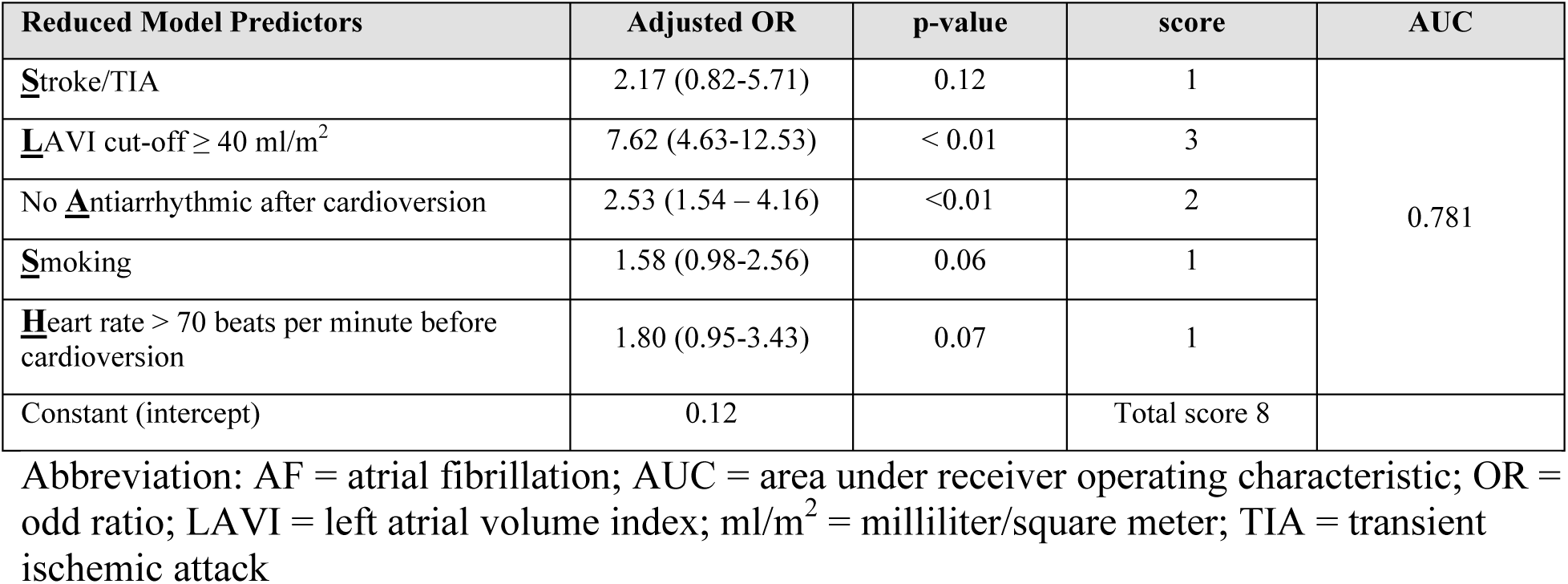
Variables included in the derivation of SLASH score prediction model.

**Table 4.** Diagnostic performance of prediction tools in predicting atrial fibrillation recurrence 6 months after external electrical cardioversion.

| SLAC score categories | Odd ratio for AF recurrence (95% CI) | Sensitivity (95% CI) | Specificity (95% CI) | Positive predictive value (95% CI) | Negative predictive value (95% CI) | LHR+ (95% CI) | LHR- (95% CI) | AUC (95% CI) |
| --- | --- | --- | --- | --- | --- | --- | --- | --- |
| Original SLAC score |  |  |  |  |  |  |  |  |
| Low risk (0-2)<br>Predicting sinus rhythm |  | 43.7%<br>(36.1-51.6) | 92.7%<br>(88.1-96.0) | 83.9%<br>(74.5-90.9) | 65.4%<br>(59.5-71.1) | 5.99<br>(3.52-10.21) | 0.61<br>(0.53-0.70) | 0.70 |
| Low risk (0-2) | 0.19 (0.12-0.30) | 21.6%<br>(16.1-28.1) | 40.7%<br>(33.2-48.6) | 29.8%<br>(22.4-38.1) | 30.9%<br>(24.9-37.5) | 0.37<br>(0.27-0.49) | 1.92<br>(1.58-2.34) |  |
| Moderate risk (3-7) | 2.56 (1.66-3.94) | 53.6%<br>(46.3-60.8) | 68.9%<br>(61.2-75.8) | 66.7%<br>(58.7-74.0) | 56.1%<br>(49.0-63.0) | 1.72<br>(1.33-2.23) | 0.67<br>(0.56-0.66) |  |
| High risk (8-14) | 3.10 (1.69-5.71) | 25.3%<br>(19.3-32.0) | 89.2%<br>(83.5-93.5) | 73.1%<br>(60.9-83.2) | 50.7%<br>(44.8-56.5) | 2.34<br>(1.42-3.86) | 0.84<br>(0.76-0.92) |  |
| ATLAS score |  |  |  |  |  |  |  |  |
| Low risk (< 6) | 0.58 (0.38-0.88) | 46.3%<br>(38.3-54.3) | 40.3%<br>(33.5-47.4) | 38.1%<br>(31.3-45.4) | 48.5%<br>(40.7-56.3) | 0.77<br>(0.63-0.95) | 1.33<br>(1.07-1.66) | 0.57 |
| Moderate risk (6-10) | 1.70 (1.06-2.73) | 59.7%<br>(52.6-66.5) | 53.8%<br>(45.7-61.7) | 61.9%<br>(54.6-68.7) | 51.5%<br>(43.7-59.3) | 1.29<br>(1.05-1.58) | 0.75<br>(0.60-0.94) |  |
| High risk (>10) | 1.76 (1.02-3.05) | 60.3%<br>(48.5-71.2) | 48.1%<br>(42.1-54.0) | 24.2%<br>(18.4-30.9) | 81.4%<br>(74.7-87.0) | 1.16<br>(0.94-1.43) | 0.83<br>(0.61-1.12) |  |
| SLASH score |  |  |  |  |  |  |  |  |
| Low risk (0-2)<br>predicting sinus rhythm |  | 43.7%<br>(36.1-51.6) | 92.7%<br>(88.1-96.0) | 83.9%<br>(74.5-90.9) | 65.4%<br>(59.5-71.1) | 5.99<br>(3.52-10.21) | 0.61<br>(0.53-0.70) | 0.74 |
| Low risk (0-2) | 0.10 (0.05-0.19) | 7.3%<br>(4.0-11.9) | 56.3%<br>(48.4-63.9) | 16.1%<br>(9.1-25.5) | 34.6%<br>(28.9-40.5) | 0.17<br>(0.10-0.28) | 1.65<br>(1.43-1.89) |  |
| Moderate risk (3-5) | 7.40 (3.88-14.10) | 54.7%<br>(47.4-61.9) | 55.7%<br>(47.8-63.4) | 58.7%<br>(51.1-66.0) | 51.7%<br>(44.1-59.2) | 1.23<br>(1.00-1.53) | 0.81<br>(0.66-1.00) |  |
| High risk (6-8) | 19.03 (8.94-40.54) | 38.0%<br>(31.1-45.3) | 88.0%<br>(82.1-92.5) | 78.5%<br>(68.8-1-86.3) | 55.3%<br>(49.1-61.3) | 3.17<br>(2.03-4.97) | 0.39<br>(0.26-0.58) |  |
| Simplified SLAC score cut-off |  |  |  |  |  |  |  |  |
| SLAC score ≥ 6 | 6.37 (4.02-10.09) | 72.1 %<br>(65.5-78.1) | 71.2%<br>(63.4-78.3) | 77.3%<br>(70.8-83.0) | 65.3%<br>(57.5-72.5) | 2.51<br>(1.93-3.26) | 0.39<br>(0.31-0.50) | 0.72<br>(0.67-0.76) |
| SLAC score ≥ 7 | 3.41 (2.17-5.35) | 71.3%<br>(63.2-78.6) | 57.8%<br>(50.9-64.4) | 52.6%<br>(45.3-59.8) | 75.4%<br>(68.2-81.8) | 1.69<br>(1.4-2.04) | 0.50<br>(0.37-0.66) | 0.65<br>(0.60-0.70) |
| SLASH score ≥ 4 | 7.65 (4.69-12.49) | 83.3 %<br>(77.3-88.3) | 60.5%<br>(52.6-67.9) | 75.9%<br>(7.8-82.9) | 70.8%<br>(64.4-76.6) | 2.11<br>(1.73-2.) | 0.28<br>(0.20-0.39) | 0.72<br>(0.67-0.76) |

Abbreviation: 95%CI = 95% confidence interval; AF = atrial fibrillation; AUC = area under receiver operating characteristic; LHR+ = positive likelihood ratio; SR = sinus rhythm

Finally, SLASH (AUC 0.781) significantly outperformed SLAC (AUC 0.705) and ATLAS (AUC 0.616) to predict AF recurrence at 6 months (p <0.01) as shown in Figure 2.

**Figure 2.**
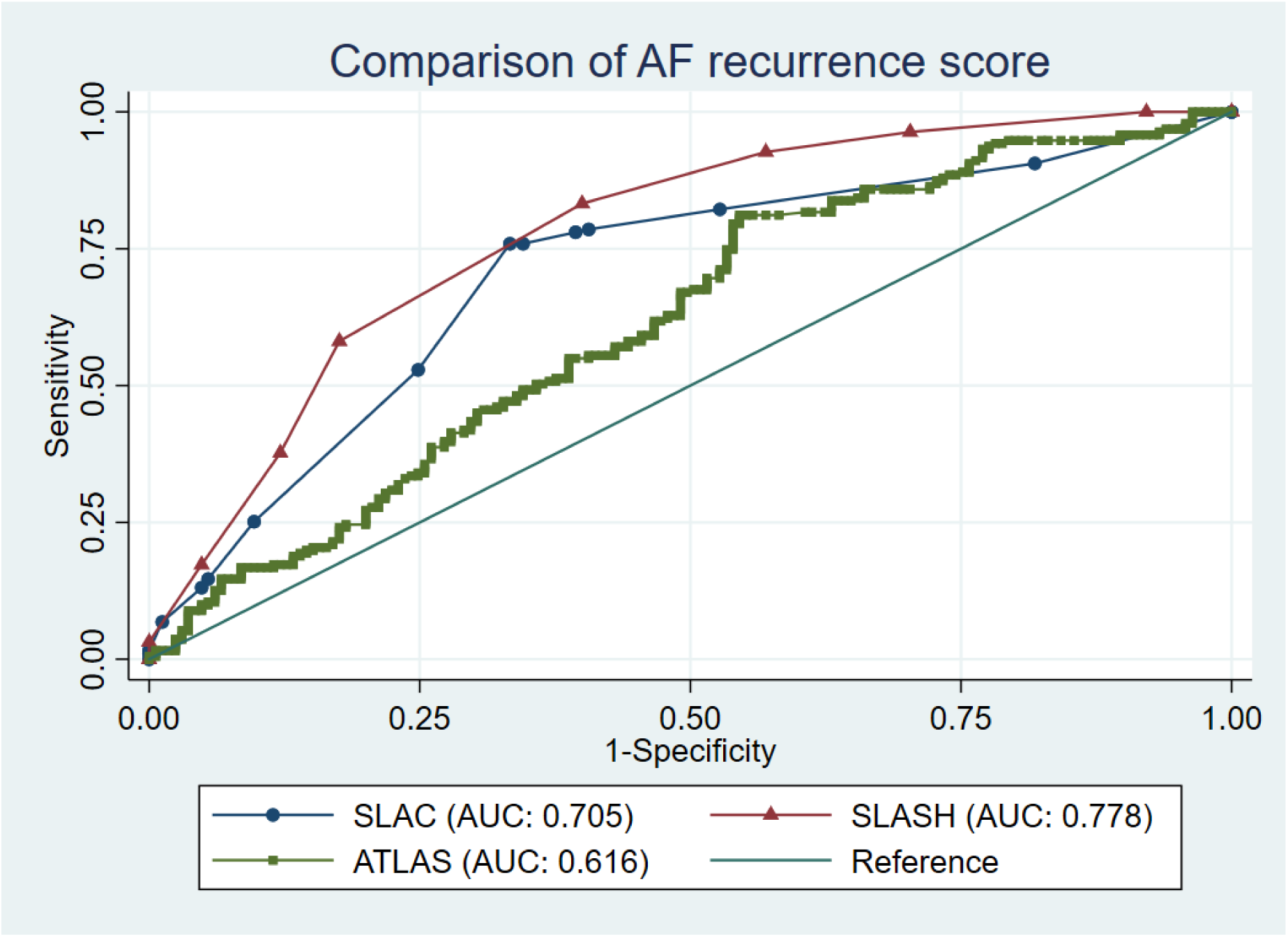
Comparison of area under receiver operative characteristic of the atrial fibrillation recurrence prediction scores Abbreviation: AF = atrial fibrillation; AUC = area under receiver operative characteristic

## Discussion

Our study addressed an often-encountered clinical dilemma post cardioversion: predicting the probability of AF recurrence. The SLAC score remains the only scoring system specifically developed to predict AF recurrence after electrical cardioversion. Other scoring models, including APPLE, ATLAS, and CAAP-AF were designed to predict AF recurrence after ablation, while the 2-point score incorporates pharmacological cardioversion (11–14). In this study, we validated the SLAC score in a larger cohort of 361 patients compared with 275 in the original derivation and 283 in the prior validation cohort (6, 7).

Electrical cardioversion is a common procedure performed in patients with AF with the goal of rhythm control. Though acutely successful in a large majority, recurrence of AF after electrical cardioversion is common. In our study, the incidence of AF recurrence at 6 months was 53.7%, closely aligning with the original study (57.5%). Multiple studies have reported AF recurrence rates ranging from 41%-52% after cardioversion. (6, 7, 15, 16).

The previously published SLAC score demonstrated fair external discrimination with an AUC of 0.704 (per-point) and 0.701 (three-group model), similar to the original study (AUC 0.765 and 0.739, respectively) (6). In our current external validation study, SLAC score demonstrated similar discrimination with an AUC of 0.704 (per-point) and 0.701 (three-group model). When individual score components were assessed, LAVI and prior stroke/TIA remained strong predictors of recurrence, while AF type and prior cardioversion were not significant. It is important to note a few differences in baseline characteristics between the derivation cohort and the current cohort. Patients in the AF recurrence group were more likely to be female and have a history of smoking, which were not significantly different in original cohort. The rates of prior stroke were two-fold higher than both the derivation and prior validation cohorts. Importantly, overall AAD use was more common among patients in the sinus rhythm maintenance group. These findings reinforce the role of AAD therapy to prevent AF recurrence.

We externally validated the diagnostic performance of the SLAC score in the current study. The SLAC score demonstrated fair but similar discrimination with AUC of 0.701 vs. 0.739 compared to the original study (6). Low SLAC scores were particularly effective in identifying patients likely to maintain sinus rhythm, with a strong positive likelihood ratio (LHR+ 5.99), whereas very high SLAC scores were specific but less sensitive for predicting recurrence. A threshold of 6 offered the best balance, with an AUC of 0.72, exceeding that of a prior study that used a score cut-off of 2 (AUC 0.602) (7).

Our secondary aim was to develop a new prediction model for AF recurrence after ECV. We found several variables that demonstrated association with AF recurrence in the univariate analysis. A few variables maintained a significant association in the multivariate model, including LAVI, lack of AAD, and a resting heart rate greater than 70 bpm. Smoking has also been included in other AF recurrence models, such as ATLAS score. AAD use was originally excluded from SLAC on the basis that it is modifiable (12). In our cohort, however, its consistent predictive value justified inclusion, as recurrence despite AAD may highlight patients who warrant escalation of care, such as earlier ablation referral.

Our updated prediction model retained two original SLAC variables (LAVI >40 ml/m² and prior stroke/TIA) and incorporated resting heart rate >70 bpm, smoking history, and post-cardioversion AAD use. The SLASH score demonstrated improved discrimination compared with the SLAC score, with an AUC of 0.78 vs. (per-point) and 0.74 vs. (three-group model), respectively. In head-to-head comparisons, the SLASH score outperformed both the SLAC and ATLAS scores. The ATLAS score also incorporates LAVI and smoking as variables similar to the updated prediction model. However, the ATLAS score was designed to predict AF recurrence after ablation and is not validated to predict AF recurrence after ECV. (12). The underperformance of the ATLAS score is likely due to the difference in the population studied. Ablation for AF is more durable with a much lower rate of recurrence. The recurrence rates of 27% in the ATLAS study are much lower than the rates in our current study, where over 50% of patients experienced recurrence (12).

Both the SLAC and SLASH scores provide clinically useful discrimination for AF recurrence after cardioversion. Although the SLAC score remains a simple and validated tool, the SLASH score provides enhanced accuracy by incorporating additional clinical variables. Together, the AF recurrence models post-ECV may support individualized treatment strategies, including AAD selection, timing of electrophysiology referral, and consideration of ablation.

Prospective studies are warranted to validate the SLASH score and determine whether score-guided management improves sinus rhythm maintenance and clinical outcomes in patients undergoing ECV for AF.

## Limitation

This study has several limitations. First, its retrospective observational design is subject to inherent bias and residual confounding. A substantial number of patients were excluded due to incomplete data, particularly the lack of an echocardiogram within the specified time frame peri-cardioversion. Second, asymptomatic or paroxysmal AF may have been underdiagnosed for AF recurrence during follow-up, potentially underestimating recurrence rates; however, our recurrence rates are comparable to prior studies. Third, recurrence was defined by ECG documentation at the time of symptoms, which may have missed short, asymptomatic episodes. Finally, the study cohort consisted primarily of Caucasian patients, which limits its generalizability to more diverse populations.

## Conclusion

The SLAC score provides fair discrimination for predicting AF recurrence at 6-months after cardioversion and reliably identifies patients likely to maintain sinus rhythm. The new SLASH score prediction model, incorporating LAVI ≥40 ml/m², prior stroke/TIA, heart rate >70 bpm, smoking history, and post-cardioversion antiarrhythmic use, demonstrated improved predictive performance and may better guide individualized rhythm management strategies. Prospective validation in diverse populations is warranted to confirm its clinical utility.

## Data Availability

most data is available in the main manuscript and supplement.

## Abbreviations

AF: atrial fibrillation
AUC: area under receiver operating characteristic
CI: confidence interval
ECV: external electrical cardioversion
LA: left atrium
LAVI: left atrial volume index
LVEF: left ventricular ejection fraction
OR: odd ratio
SR: sinus rhythm
TIA: transient ischemic attack

---

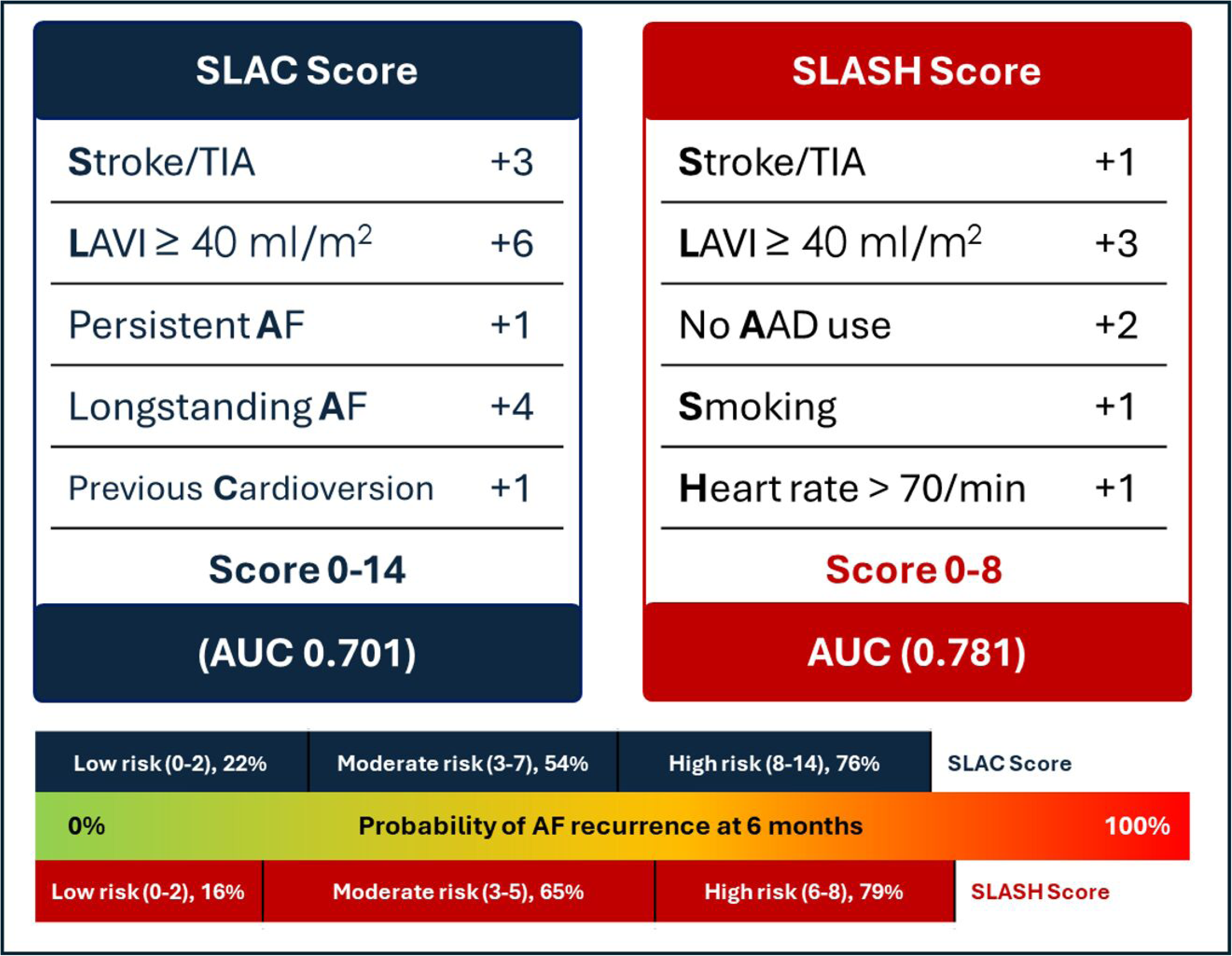

## References

1. Kirchhof P, Camm AJ, Goette A, Brandes A, Eckardt L, Elvan A, et al. Early Rhythm-Control Therapy in Patients with Atrial Fibrillation. The New England journal of medicine. 2020;383(14):1305–16.

2. Sandhu RK, Smigorowsky M, Lockwood E, Savu A, Kaul P, McAlister FA. Impact of Electrical Cardioversion on Quality of Life for the Treatment of Atrial Fibrillation. The Canadian journal of cardiology. 2017;33(4):450–5.

3. Gilbert KA, Hogarth AJ, MacDonald W, Lewis NT, Tan LB, Tayebjee MH. Restoration of sinus rhythm results in early and late improvements in the functional reserve of the heart following direct current cardioversion of persistent AF: FRESH-AF. Int J Cardiol. 2015;199:121–5.

4. Qeska D, Qiu F, Manoragavan R, Wijeysundera HC, Cheung CC. Relationship between wait times and postatrial fibrillation ablation outcomes: A population-based study. Heart rhythm. 2024;21(9):1477–84.

5. Ecker V, Knoery C, Rushworth G, Rudd I, Ortner A, Begley D, et al. A review of factors associated with maintenance of sinus rhythm after elective electrical cardioversion for atrial fibrillation. Clinical cardiology. 2018;41(6):862–70.

6. Thangjui S, Yodsuwan R, Thyagaturu H, Navaravong L, Zoltick J. A Prognostic Score To Predict Atrial fibrillation Recurrence After External Electrical Cardioversion-SLAC Score. Crit Pathw Cardiol. 2022;21(4):194–200.

7. Águila-Gordo D, Jiménez-Díaz J, Negreira-Caamaño M, Martínez-Del Rio J, Ruiz-Pastor C, Sánchez Pérez I, et al. Usefulness of risk scores and predictors of atrial fibrillation recurrence after elective electrical cardioversion. Annals of noninvasive electrocardiology : the official journal of the International Society for Holter and Noninvasive Electrocardiology, Inc. 2024;29(1):e13095.

8. von Elm E, Altman DG, Egger M, Pocock SJ, Gøtzsche PC, Vandenbroucke JP. The Strengthening the Reporting of Observational Studies in Epidemiology (STROBE) statement: guidelines for reporting observational studies. Journal of clinical epidemiology. 2008;61(4):344–9.

9. Lang RM, Badano LP, Mor-Avi V, Afilalo J, Armstrong A, Ernande L, et al. Recommendations for cardiac chamber quantification by echocardiography in adults: an update from the American Society of Echocardiography and the European Association of Cardiovascular Imaging. Journal of the American Society of Echocardiography : official publication of the American Society of Echocardiography. 2015;28(1):1–39.e14.

10. Collins GS, Reitsma JB, Altman DG, Moons KGM. Transparent reporting of a multivariable prediction model for individual prognosis or diagnosis (TRIPOD): the TRIPOD Statement. BMC Medicine. 2015;13(1):1.

11. Kornej J, Hindricks G, Shoemaker MB, Husser D, Arya A, Sommer P, et al. The APPLE score: a novel and simple score for the prediction of rhythm outcomes after catheter ablation of atrial fibrillation. Clinical research in cardiology : official journal of the German Cardiac Society. 2015;104(10):871–6.

12. Mesquita J, Ferreira AM, Cavaco D, Moscoso Costa F, Carmo P, Marques H, et al. Development and validation of a risk score for predicting atrial fibrillation recurrence after a first catheter ablation procedure - ATLAS score. Europace : European pacing, arrhythmias, and cardiac electrophysiology : journal of the working groups on cardiac pacing, arrhythmias, and cardiac cellular electrophysiology of the European Society of Cardiology. 2018;20(Fi_3):f428–f35.

13. Winkle RA, Jarman JW, Mead RH, Engel G, Kong MH, Fleming W, et al. Predicting atrial fibrillation ablation outcome: The CAAP-AF score. Heart rhythm. 2016;13(11):2119–25.

14. Fornengo C, Antolini M, Frea S, Gallo C, Grosso Marra W, Morello M, et al. Prediction of atrial fibrillation recurrence after cardioversion in patients with left-atrial dilation. European Heart Journal - Cardiovascular Imaging. 2014;16(3):335–41.

15. Okçün B, Yigit Z, Küçükoglu MS, Mutlu H, Sansoy V, Güzelsoy D, et al. Predictors for maintenance of sinus rhythm after cardioversion in patients with nonvalvular atrial fibrillation. Echocardiography (Mount Kisco, NY). 2002;19(5):351–7.

16. Disertori M, Lombardi F, Barlera S, Latini R, Maggioni AP, Zeni P, et al. Clinical predictors of atrial fibrillation recurrence in the Gruppo Italiano per lo Studio della Sopravvivenza nell’Infarto Miocardico-Atrial Fibrillation (GISSI-AF) trial. American heart journal. 2010;159(5):857–63.

